# Quantifying the success of measles vaccination campaigns in the Rohingya refugee camps

**DOI:** 10.1101/19008417

**Authors:** Taylor Chin, Caroline O. Buckee, Ayesha S. Mahmud

## Abstract

In the wake of the Rohingya population’s mass migration from Myanmar, one of the world’s largest refugee settlements was constructed in Cox’s Bazar, Bangladesh to accommodate nearly 900,000 new refugees. Refugee populations are particularly vulnerable to infectious disease outbreaks due to many population and environmental factors. A large measles outbreak, with over 2,500 cases, occurred among the Rohingya population between September and December 2017. Here, we estimate key epidemiological parameters and use a dynamic mathematical model of measles transmission to evaluate the effectiveness of the reactive vaccination campaigns in the refugee camps. We also estimate the potential for subsequent outbreaks under different vaccination coverage scenarios. Our modeling results highlight the success of the vaccination campaigns in rapidly curbing transmission and emphasize the public health importance of maintaining high levels of vaccination in this population, where high birth rates and historically low vaccination coverage rates create suitable conditions for future measles outbreaks.

## 1 Background

Since violence in Myanmar intensified in August 2017, nearly 900,000 Rohingya refugees have migrated from the Rakhine State of Myanmar and settled in refugee settlements in Cox’s Bazar, Bangladesh [1]. Mass population displacement has been connected to adverse public health outcomes due to a plethora of risk factors, including heightened insecurity, reduced healthcare access, and inadequate living conditions [2]. Despite global gains in reducing the burden of vaccine-preventable infectious diseases [3], low vaccination coverage rates and overcrowding conditions in refugee camps often give rise to large-scale outbreaks in these settings [4–8].

Given the crowded conditions in the refugee camps and the low rates of measles immunization among the Rohingya population, a large measles outbreak broke out in the Rohingya refugee camps in Cox’s Bazar in late 2017, with over 2,500 cumulative suspected cases of measles reported from September to December 2017 [9–11]. Measles is a highly contagious respiratory infection caused by a paramyxovirus [12]. Transmission occurs through respiratory droplets or direct contact with secretions from the nose or throat of infected individuals [13]. Two vaccination campaigns were conducted in response to the outbreak; the first campaign ran from September 16, 2017 to October 4, 2017 and delivered the measles and rubella (MR) vaccine to 135,519 children aged 6 months to 15 years old [14]. A second vaccination campaign ran from November 18, 2017 to December 2, 2017 and vaccinated 323,940 children in the same age group [15].

We first evaluate the success of these vaccination campaigns in curbing the measles outbreak in the camps, by quantifying changes in the effective reproductive number (R_e_) and estimating the number of cases averted. Our results highlight the success of the reactive vaccination campaigns—a remarkable public health achievement given the very low rates of prior immunization among the Rohingya population.

We find that although these campaigns achieved high coverage, future measles outbreaks are still likely. First, the Rohingya population has historically low vaccination coverage rates; a survey conducted in March 2018 found that 42.9% of children under the age of four had not received even a single dose of an injectable vaccine in Myanmar, and only 2.8% of them had received five or more doses [16]. Second, the birth rate among the refugee population is high, which rapidly replenishes the susceptible population. UNICEF estimates that approximately 60 Rohingya babies were born in Cox’s Bazar each day in the nine months following the initial wave of migration in August 2017 [17]. We use a mathematical model to estimate the potential for subsequent measles outbreaks under different vaccination coverage scenarios. Our findings have important public health implications, as greater understanding of the dynamics of measles in Cox’s Bazar can help inform future resource prioritization and vaccination targets.

## 2 Methods

Daily case data was obtained from the World Health Organization’s (WHO) reported cases of measles and rubella in Cox’s Bazar from September 6, 2017 to November 25, 2017 [9]. The data include 1,708 confirmed and suspected cases of measles and rubella during this time period. All suspected cases are included in this analysis given the disease’s characteristic rash [12], which facilitates syndromic reporting of measles. During an early blood sampling of 89 suspected cases of measles during the outbreak, only 1% of samples tested positive for rubella-specific immunoglobulin M (IgM) [9]. The suspected cases are therefore assumed to be measles rather than rubella. We also assume that all suspected cases occurred among the refugee population given the WHO’s finding that only 1% of cases occurred among the host community population [9].

### 2.1 Estimation of R_e_

We first estimated the daily effective reproductive number for the measles outbreak, in order to estimate the basic reproductive number—a key epidemiological parameter. The basic reproductive number is defined as the expected number of secondary cases from a typical infectious case in a completely susceptible host population [18]. The reproductive number quantifies the transmissibility of an infection, thereby informing key attributes such as the herd immunity threshold.

We estimated the effective reproductive number at time *t* using the approach developed by Wallinga and Teunis [19], where knowledge of the serial interval distribution and incident cases is used in a likelihood-based estimation procedure that considers pairs of cases. The serial interval distribution of measles used in this analysis was based on a meta-analysis of existing studies of measles, and was assumed to be normally distributed with a mean of 11.9 days and standard deviation of 2.6 days [20]. Table 1 reports all parameters used in this analysis and their sources. The *EpiEstim* package in *R* was used to estimate the daily effective reproductive number over the course of the outbreak. The 95% credible intervals were estimated using 1,000 simulations and 14-day time windows to increase precision in estimates of the daily effective reproductive number [21].

**Table 1:** Summary of Input Parameters.

| Parameter | Value/Distribution | Notes | Source |
| --- | --- | --- | --- |
| Estimation of $R_e$ | | | |
| Measles serial interval distribution | $N(11.9, 2.6^2)$ | An estimated serial interval distribution reported from a household transmission study from Providence, R.I. by Chapin (1925) | Vink, Bootsma, and Wallinga [20] |
| Susceptible population at start of 2017 outbreak (%), $s_0$ | Triangle(0.05,0.25,0.15) | Given the low vaccination rates in Myanmar [16], we assumed that most Rohingya adults were already immune to measles due to a resultant low average age of first infection. This assumption seems reasonable given that 83% of suspected cases were among children < 5 years old, and 96% of suspected cases were among children < 15 years old during the 2017 outbreak [9]. Approximately 50% of the Rohingya population in Cox's Bazar is < 15 years old [16,22]. 72% of measles cases in the 2017 outbreak as of November 18, 2017 had no history of measles vaccination [9]. | Bhatia *et al.* [16], WHO [9], MSF [22] |
| Mechanistic Measles Model |  |  |  |
| Basic reproductive number, $R_0$ | Gamma(shape=16.2, rate=1.2) | Fit gamma distribution to histogram of $R_0$ values, which were estimated in the analysis using the relationship $R_e = R_0 * s_0$ | Estimated in analysis |
| Rohingya population size, December 2017 | 579,661 | The total influx of Rohingya into Cox's Bazar from August 25, 2017 to December 5, 2017, excluding those that settled in the host community | WHO [23] |
| Rohingya population size, April 2019 | 870,534 | The number of Rohingya refugees identified in the refugee camps as of April 2019, excluding those registered before August 31, 2017 | ISCG [1] |
| Birth rate (/10,000/day), $\mu$ | 1.45 | Save the Children estimated that 48,000 Rohingya children would be born in Cox's Bazar in 2018 [24], which is in line with UNFPA's estimate from December 2017 that 10,000 Rohingya women would give birth from January to March 2018 [25]. | Save the Children [24], UNFPA [25] |
| Death rate (/10,000/day), $\mu$ | 1.45 | Assumed to be the same as the birth rate | Assumption |
| Recovery rate (days<sup>-1</sup>), $\gamma$ | 0.11 | 1/average duration of infectiousness (9 days) | McLean *et al.* [26] |
| Transmission coefficient (days<sup>-1</sup>), $\beta$ | — | $R_0 / (\text{average duration of infectiousness} * \text{population size})$ | Estimated in analysis |
| Susceptible population at start of next outbreak (%), $s_U$ | Uniform(0.05,0.15) | 349,603 children < 15 years old were targeted for second vaccination campaign [9].<br>If 135,519 children were vaccinated in the first round and 323,940 in the second round, 38.8% (135,519/349,603) and 92.7% (323,940/349,603) of children were vaccinated in these campaigns, respectively<br>Use binomial calculations to estimate probability of child receiving 0, 1, and 2 MR doses as 4.5%, 59.6% and 35.9%, respectively<br>If 4.5% of children received 0 doses, approximately 15,715 children were not vaccinated by the end of the vaccination campaigns<br>Add estimated 72,000 births (48,000*1.5) that have occurred since the vaccination campaigns; annual estimate of 48,000 is the same estimate from Save the Children [24] that informs the birth rate<br>Estimate current proportion of children susceptible as $(15,715 + 72,000) / 870,534 = 0.10$ to inform the middle of the uniform distribution range | Assumption |

### 2.2 Mechanistic Measles Model

Based on our estimated R_e_, we estimated the basic reproductive number, R_0_, in order to simulate the dynamics of measles using a mechanistic transmission model. To account for uncertainty at the start of outbreak, we used a range of effective reproductive numbers, R_e_ to estimate R_0_ according to the relationship *R*_e_ = *R*_0_ *\* s*_0_, where s_0_ is the fraction of the population susceptible at the start of the outbreak and R_e_ is the effective reproductive number at the start of the epidemic. The R_e_ range was chosen based on when the epidemic curve suggests the measles outbreak took off. Effective reproductive numbers between 1.4 and 3.1 correspond to values calculated using 14-day intervals that start between days 6 and 16 into the outbreak. Here, we assume mass-action and that the population is well-mixed [18]. Latin Hypercube Sampling (LHS) was used to capture uncertainty around parameter estimates by efficiently sampling from the joint distribution of parameter inputs [27]. Using this method, 1,000 values of s_0_ and R_e_ were sampled from their assumed probability distributions (Table 1). A gamma distribution was fit to the resultant 1,000 estimated values of R_0_. The median and 2.5% and 97.5% percentile of the reproductive number are reported, and a histogram of estimated values is shown to reflect estimates based on the LHS procedure (Supplementary Figure 1).

A deterministic, Susceptible-Infectious-Recovered (SIR) model, which incorporates birth and death rates, was used to model the size of measles outbreaks in the refugee camps under the scenario in which reactive vaccination had not been implemented in 2017 and under different vaccination scenarios for the next outbreak (Figure 1). The model assumes random mixing in the population and does not consider age-dependent heterogeneity in transmission that occurs in other settings [28].

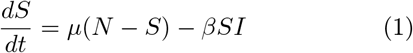

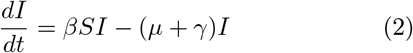

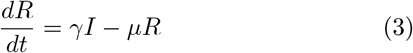

**Figure 1:**
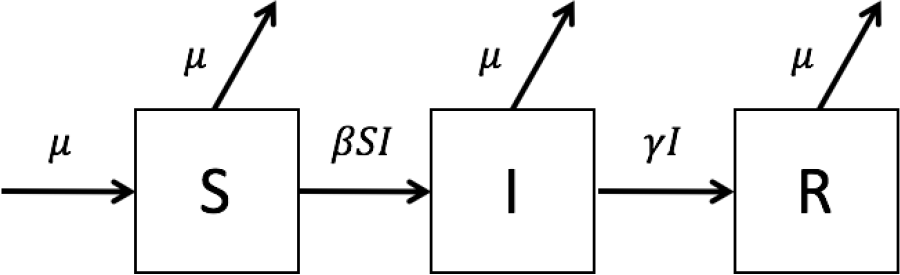
Schematic representation of disease states in a model of a measles outbreak in Cox’s Bazar. Individuals are either susceptible to measles (S), infectious with measles (I), or recovered from and immune to measles infection (R), either due natural immunity from infection or prior vaccination. *β* represents the transmission coefficient, *γ* is the recovery rate, and *µ* is the birth and death rate. The model starts with one infected (and infectious) measles case (*i*.*e*., I(0) = 1). The number of recovered individuals at time = 0 was calculated as R(0) = *N* − *S*(0) − *I*(0). The model tracks the number of individuals that move between the compartments each day and is run for one year using the ordinary differential equations 1-3.

The transmission coefficient, *β*, was estimated according to the relationship *β* = *R*_0_*/ND*, where *N* is the total population (*i*.*e*., S + I + R) and *D* is the average duration of infectiousness (Table 1). A measles-specific death rate was not included in the model since it was assumed to be negligible, as there were only 3 measles-attributed deaths from August 25, 2017 to December 9, 2017 [15, 29]. Immunity from both natural infection and vaccination was assumed to be life-long. Two doses of the MMR vaccine have been shown to confer immunity for at least 20 years [30]. The model uses one infected case as the initial value for the number of individuals in the infected compartment, I(0). The initial value for the recovered compartment was calculated as R(0) = *N* − *S*(0) − *I*(0).

#### 2.2.1 Estimating the Impact of Vaccination on the 2017 Outbreak

The population included in this analysis was the 579,661 Rohingya that migrated to Cox’s Bazar after August 25, 2017 and settled in the refugee camps [23]. The proportion of the population that was estimated to be susceptible at the start of the outbreak, *s*_0_, was estimated by considering prior vaccination rates and the age distribution of the Rohingya population (Table 1). The model runs for 365 days to reflect one year following the start of the outbreak. LHS was used to take into account uncertainty in parameter estimates; 1,000 values for each parameter were sampled from the assumed parameter probability distributions (Table 1). A histogram of values for the estimated final size of the 2017 epidemic in the absence of vaccination is shown to reflect uncertainty in estimates (Supplementary Figure 2).

#### 2.2.2 Modeling the Impact of Vaccination on the Next Outbreak

To model the impact of vaccination on subsequent measles epidemics, we assumed a total Rohingya population size of 870,534 as of April 2019 [1]. The SIR model described above (Figure 1) was used to estimate the size of subsequent epidemics based on varying assumptions for vaccination coverage rates. The model incorporates infection rates during the 2017 outbreak and the vaccination coverage rates of the campaigns conducted in 2017 by reducing the proportion of the population that is susceptible (*s*) through the formula previously described by Gastañaduy *et al*. (2018) [31]

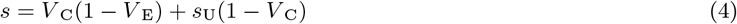

where V_C_ is *≥* 1 dose coverage of the Measles, Mumps, and Rubella (MMR) vaccine, V_E_ is the median effectiveness of 1 dose of MMR vaccine (94%) [26], and s_U_ is the proportion of unvaccinated individuals who are susceptible. This approach assumes vaccination is implemented as one-time interventions between the end of the 2017 outbreak and now. Table 1 reports all parameters and calculations used as inputs in the mathematical model. LHS was again used to take into account uncertainty in parameter estimates.

For this analysis, the SIR model was run for two years to estimate the impact of vaccination on the next outbreak. The cumulative number of cases over two years was calculated for vaccination coverage rates of 10%, 50%, and 90% of the at-risk population. Cumulative cases were estimated as the cumulative proportion of the population that moves into the infectious compartment in the SIR framework. Distributions of our estimates for the cumulative number of cases at two years under the different vaccination rate scenarios are presented. To contrast these results with the scenario of the next measles outbreak in the absence of vaccination, the same model used to estimate the size of the 2017 outbreak in the absence of vaccination was used with two changes: 1) the total population size was increased to 870,534 [1], and 2) the proportion of the population that was assumed to be susceptible at the start of the outbreak was estimated as s_u_ instead of s_0_ (Table 1) to incorporate the two vaccination campaigns conducted in 2017 and births that have occurred since December 2017.

## 3 Results

### 3.1 Quantifying changes to R_e_

During the 2017 measles outbreak in Cox’s Bazar, the total number of measles cases from September 6, 2017 to November 25, 2017 was 1,708. The number of reported measles cases reached a peak of 86 incident cases on November 18, 2017, followed by a decline in cases (Figure 2A). Current measles transmission appears to be ongoing but limited in the refugee camps, with 323 suspected cases reported from January 1, 2019 to May 4, 2019 [32].

**Figure 2:**
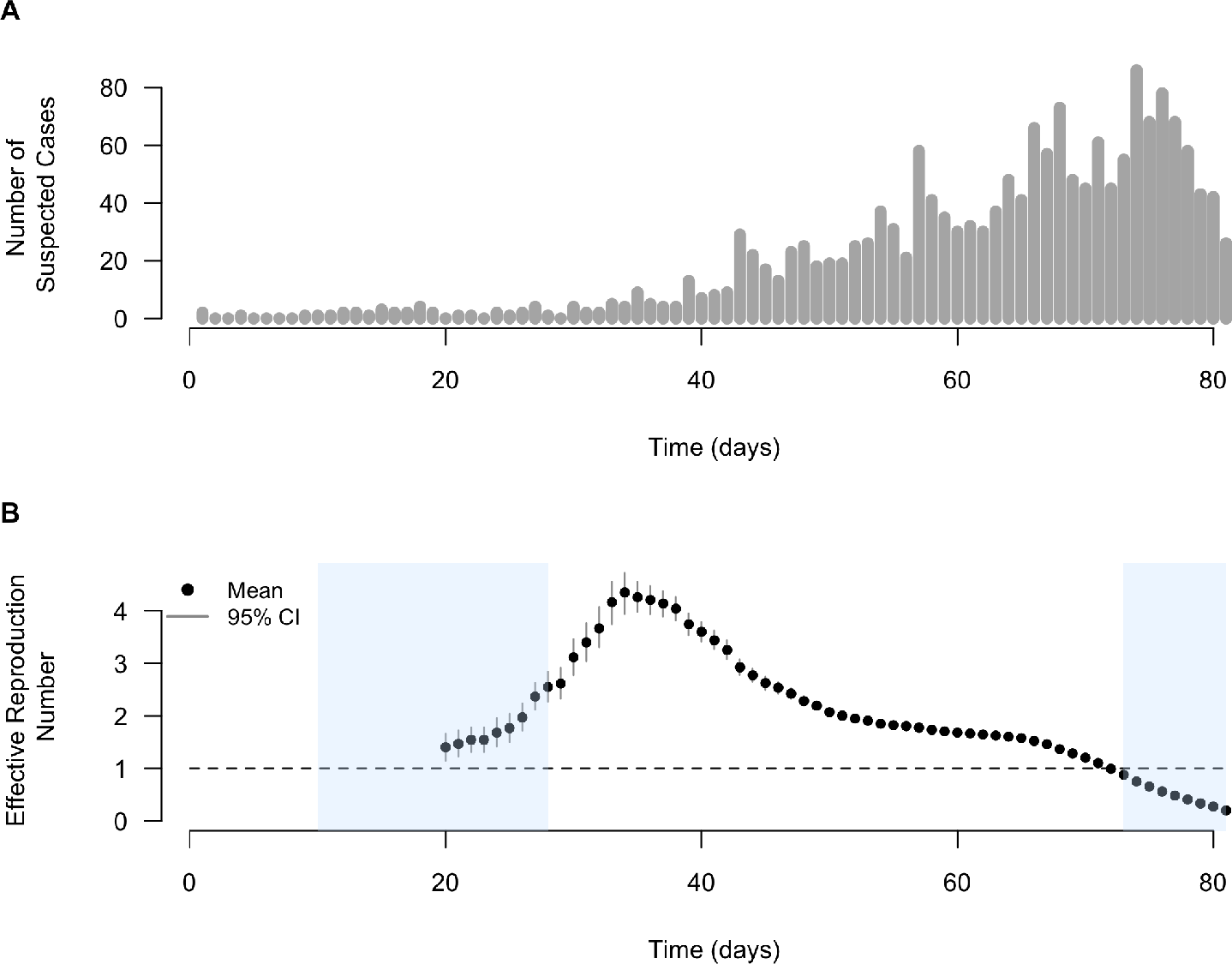
A) Confirmed and suspected measles and rubella cases in Cox’s Bazar from September 6, 2017 to November 25, 2017. B) Estimated daily effective reproductive number of measles in Cox’s Bazar in the 2017 outbreak using the Wallinga and Teunis method over sliding 14-day windows. Time scale is days since the start of the outbreak on September 6, 2017. Blue shaded areas indicate vaccination campaigns (first campaign from September 16, 2017 to October 4, 2017 and second campaign from November 18, 2017 to December 2, 2017). The 95% credible intervals were estimated using 1,000 simulations.

The daily effective reproductive number ranged from approximately 1.4 (2.5% percentile 1.1 to 97.5% percentile 1.7) at the end of the third week of the outbreak to a maximum of 4.3 (2.5% percentile 4.0 to 97.5% percentile 4.8) at the end of the fifth week, or the week ending on October 10, 2017 (Figure 2B). The first measles vaccination campaign conducted in response to the outbreak appears to have been highly successful in curbing the transmission of measles, as the daily effective reproductive number declined shortly after the end of the two-week campaign. The second vaccination campaign, meanwhile, was conducted starting in December, when the daily effective reproductive number was already below 1. By the middle of the 11th week, or mid-November, the effective reproductive number had already dipped below 1. After restricting the maximum estimated basic reproductive number to 20 based on the ranges for R_0_ commonly reported in the literature [33], the median basic reproductive number is estimated to be 13.7 with a standard deviation of 3.3 (Supplementary Figure 1).

### 3.2 Estimating the Impact of Vaccination on the 2017 Outbreak

If the two vaccination campaigns had not been implemented in response to the 2017 outbreak, our model estimates a median final epidemic size of approximately 79,404 measles cases (standard deviation (sd) of 29,073). This result reflects the high transmissibility of measles, and suggests that after one year from one infectious individual entering the Rohingya population in September 2017, in the absence of interventions, almost all susceptible individuals—or equivalently, approximately 14% of the total population—would have contracted measles. Considering that our model therefore indicates a near 100% attack rate among the susceptible population, the fact that only approximately 2,500 cumulative cases occurred from September to the end of December 2017 is a testament to the tremendous success of the vaccination campaigns that were implemented by the WHO, the Bangladesh Ministry of Health, Family and Welfare (MoHFW), and their partners in response to the measles outbreak [9–11]. Our results suggest that nearly 77,000 cases were averted as a result of the reactive vaccination campaigns conducted in late 2017 (Figure 3). The swift response of WHO, the MoHFW, and partner organizations, considering the challenging circumstances of accommodating over 650,000 new arrivals by December 2017, is a remarkable public health achievement [34].

**Figure 3:**
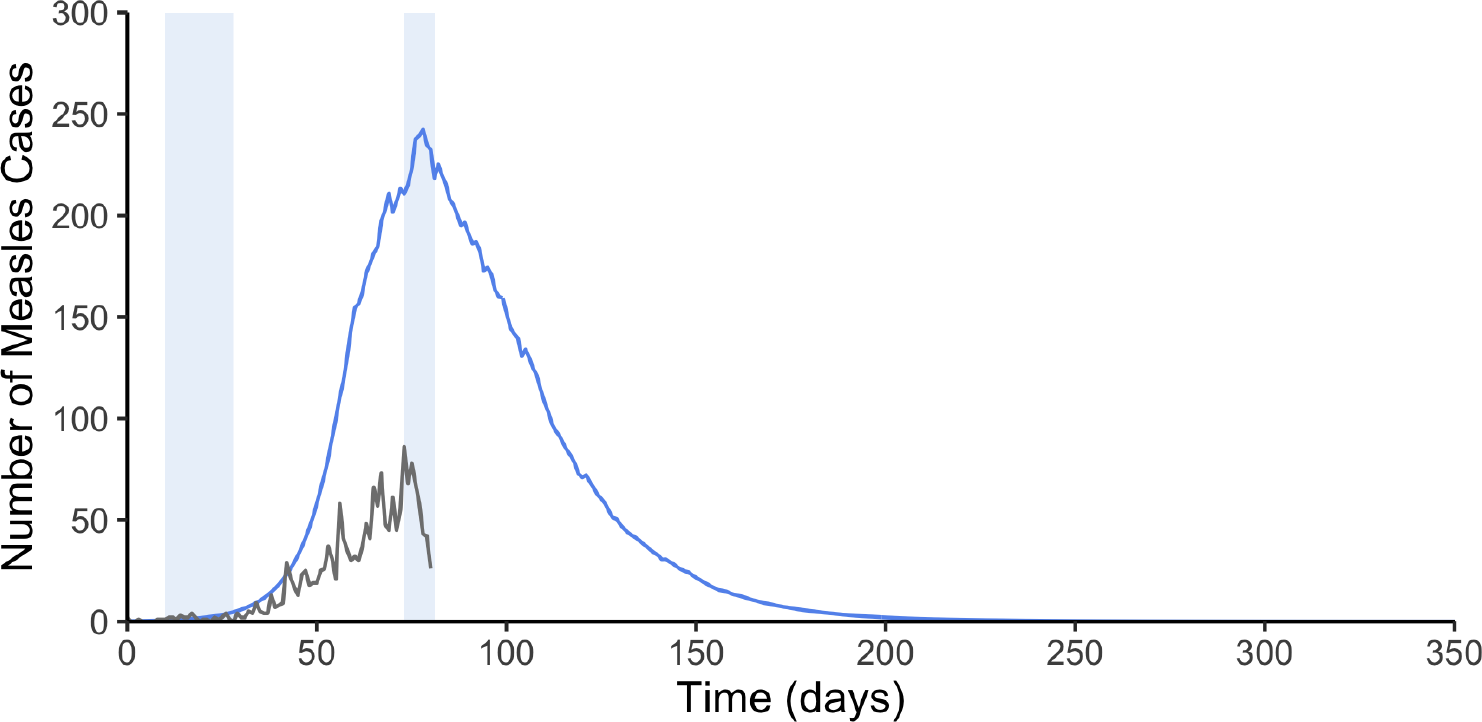
Confirmed and suspected measles and rubella cases in Cox’s Bazar from September 6, 2017 to November 25, 2017 shown in gray. Estimated median number of measles cases in the absence of vaccination over a one-year period beginning on September 6, 2017 shown in blue. The median is based on 1,000 parameter combinations using LHS. Light blue shaded areas indicate vaccination campaigns (first campaign from September 16, 2017 to October 4, 2017 and second campaign from November 18, 2017 to December 2, 2017).

### 3.3 Modeling the Impact of Vaccination on the Next Outbreak

When our model is applied to the current population size under different vaccination scenarios, the model estimates that the median number of measles cases after two years would range from approximately 74,500 cases if 90% of the population is vaccinated to 90,000 cases under the scenario of a 10% vaccination coverage rate (Figure 4A). These results contrast with the larger median estimate of approximately 93,700 cases, or 11% of the total Rohingya population, predicted for the next measles outbreak in this setting under the scenario of no vaccination (Supplementary Figure 3). Vaccination therefore has the potential to avert between approximately 4,000 and 19,000 cases over two years depending on the coverage rate. Figure 4B shows estimates in terms of boxplots to reflect the uncertainty in our model’s results due to parameter uncertainty.

**Figure 4:**
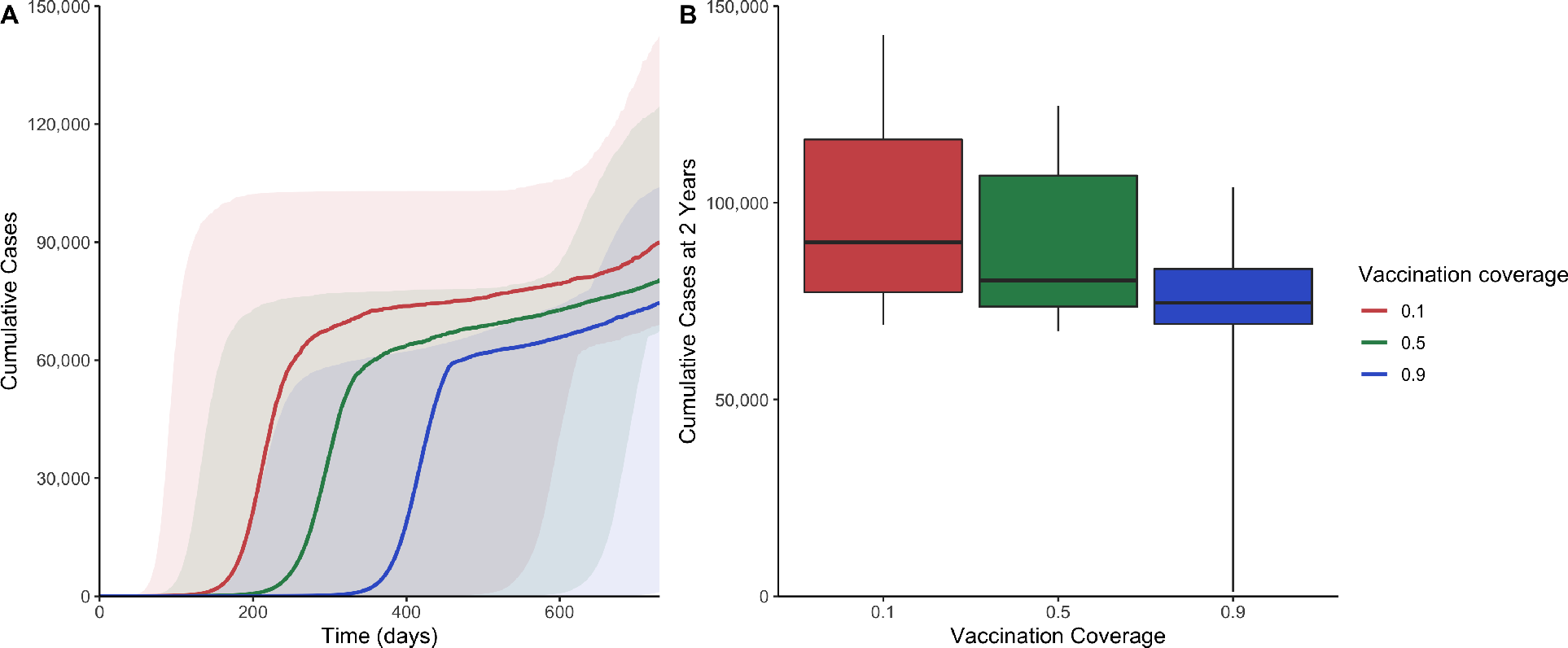
A) Cumulative number of measles cases over time under varying scenarios of vaccination coverage rates. Shaded areas represent the range between estimates’ 10% and 90% percentiles from LHS sampling of 1,000 parameter combinations B) Boxplots of the cumulative number of measles cases at 2 years (730 days) using 1,000 LHS parameter combinations under the same vaccination coverage rate scenarios. Maximum and minimum values represent 90% and 10% percentiles, respectively.

## 4 Discussion

Our analysis highlights the immense success of the two large-scale vaccination campaigns in averting measles cases during the 2017 outbreak in Cox’s Bazar and supports the importance of maintaining high vaccination coverage rates in the population going forward. Our findings on the estimate of the basic reproductive number in this setting (median = 13.7; sd = 3.3) are in line with previous estimates reported in the literature [33].

We find that nearly 77,000 cases were averted over the course of one year due to vaccination in the 2017 outbreak. The daily effective reproductive number declined from its maximum value of 4.3 at the end of week 5 to below 1 by the middle of week 11. When we apply our model to estimate the impact of vaccination on subsequent outbreaks, depending on the coverage rate in the population, vaccination could avert between 4,000 and 19,000 cases over two years. The near 100% attack rate found among susceptibles in the absence of vaccination reinforces the importance of maintaining vaccination as a key public health priority among the Rohingya community in Cox’s Bazar.

Our study has several important limitations. The estimate for the basic reproductive number may be biased due to two possible ways in which the serial interval distribution from the literature may overestimate the serial interval distribution in this specific setting. First, it is possible that other intervention measures such as contact tracing and isolation of infectious cases were implemented during the outbreak, which would have the effect of shortening the serial interval for such individuals. However, press articles or WHO reports do not mention these interventions, and it may be reasonable to assume that these interventions were not widely used given their infeasibility in this setting. Secondly, the contraction of the serial interval described by Kenah, Lipsitch, and Robins [35] in high transmission settings is not considered in this analysis. Instead, a constant serial interval distribution is assumed. For these reasons, the serial interval used in this analysis may represent an overestimation of the actual serial interval in this setting, which would result in an overestimation of the basic reproductive number given the positive correlation between the two measures. Future analyses could examine how the effective reproductive number at time *t* would change if a hazard-based estimator described by Kenah, Lipsitch, and Robins [35] was instead used in order to address potential serial interval contraction.

Second, reporting rates and how these rates changed over time are also not considered in this analysis. One possible scenario is that identification of measles was worse at the start of the epidemic relative to later in the epidemic, and that fewer measles cases were therefore reported in the early phase of the epidemic. If this were the case, the initial case counts may underestimate the true extent of measles transmission at the start of the epidemic, which would affect the calculation for the reproductive number. Assumptions about changes in the reporting rate could be incorporated in future work, such that the reported daily cases are inflated by a different fraction across time, for example using a method similar to that used by White *et al*. [36] in their investigation of the 2009 H1N1 pandemic.

Third, our results pertain to a specific outbreak among the Rohingya population in Cox’s Bazar and may not generalize to other populations. Importantly, our assumption of homogeneous mixing may not hold in other settings. Despite these limitations, this analysis provides estimates for the transmissibility of measles in the setting of Cox’s Bazar as well as quantifies the impact of vaccination. Starting in December 2017, the WHO and its partners implemented MR vaccination at border check points [37]. From February to December 2018, approximately 29,000 MR doses were delivered to children [38]. The WHO currently supports the Ministry of Health in routine measles surveillance [39]. These efforts are highly commendable and should be recognized.

Importantly, however, our findings emphasize the necessity of maintaining ongoing surveillance and of consistently implementing preventive interventions like vaccination. Recently, attention in the refugee camps has been focused primarily on diarrheal diseases in Cox’s Bazar [40]. At the same time, according to the WHO’s Early Warning, Alert, and Response System (EWARS) in Cox’s Bazar, approximately 10 to 20 cases of suspected measles or rubella have been reported to EWARS each week since the start of 2019, signaling ongoing measles transmission [41].

Against this backdrop of competing public health concerns, it will be important to sustain routine vaccination of vaccine-preventable childhood infections like measles given the high birth rate in the camps and evidence that measles transmission continues to linger. Estimates of the basic reproductive number and size of the next outbreak from this analysis reinforce the public health importance of maintaining the high vaccination coverage rates that are known to be necessary for preventing outbreaks of highly-transmissible infections such as measles.

## Data Availability

A link to the raw data is provided in the paper.

## Author Contributions

TC performed the data analyses and drafted the manuscript. COB and ASM designed the research. All authors revised the manuscript and approved the final version.

## Supplementary Material

**Figure 1:**
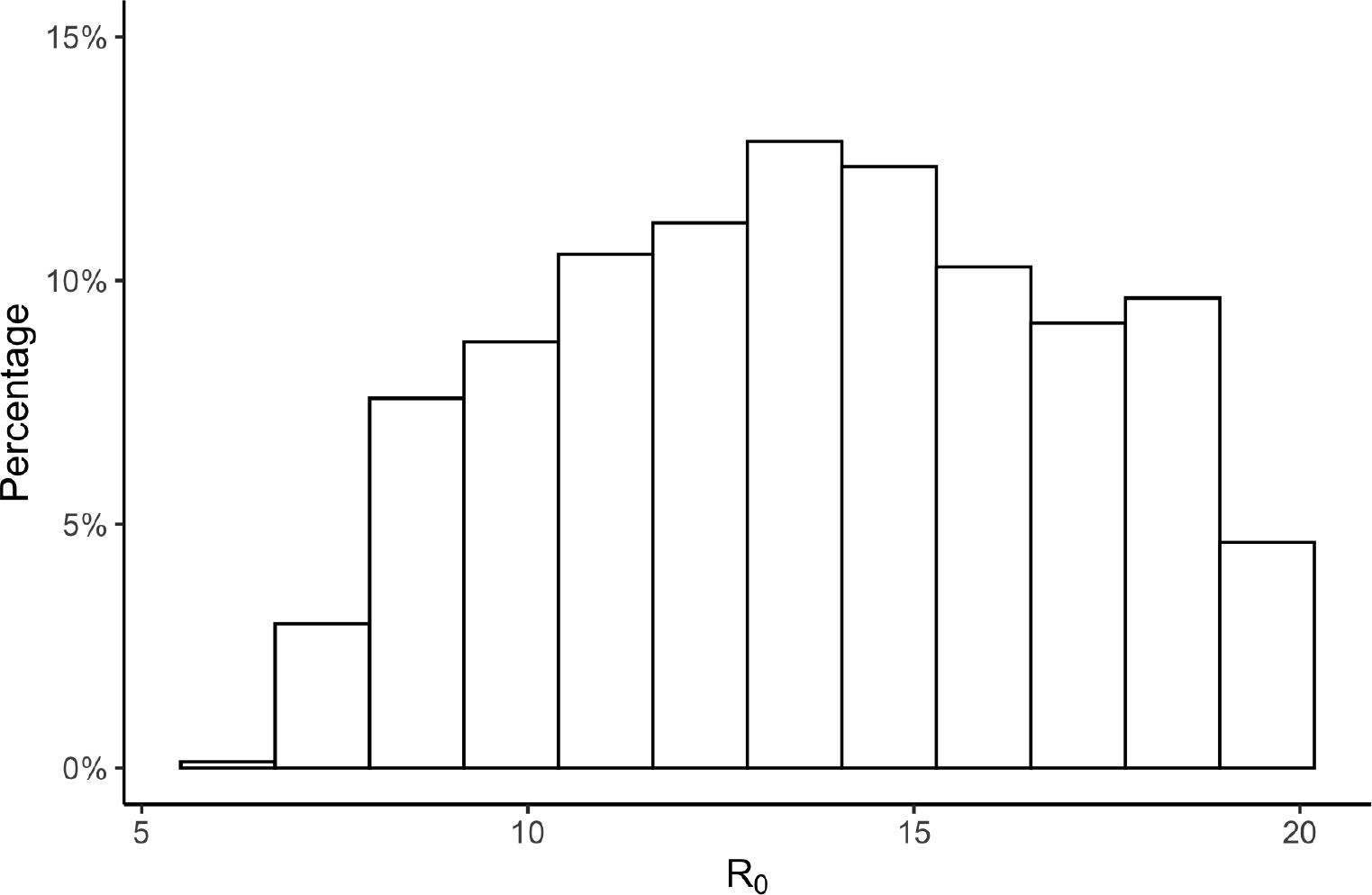
Histogram of estimated R_0_ values using LHS with 1,000 parameter combinations, where the maximum has been restricted to 20. Summary statistics: 13.67 (mean), 13.69 (median), 7.76 (2.5%) and 19.49 (97.5%)

**Figure 2:**
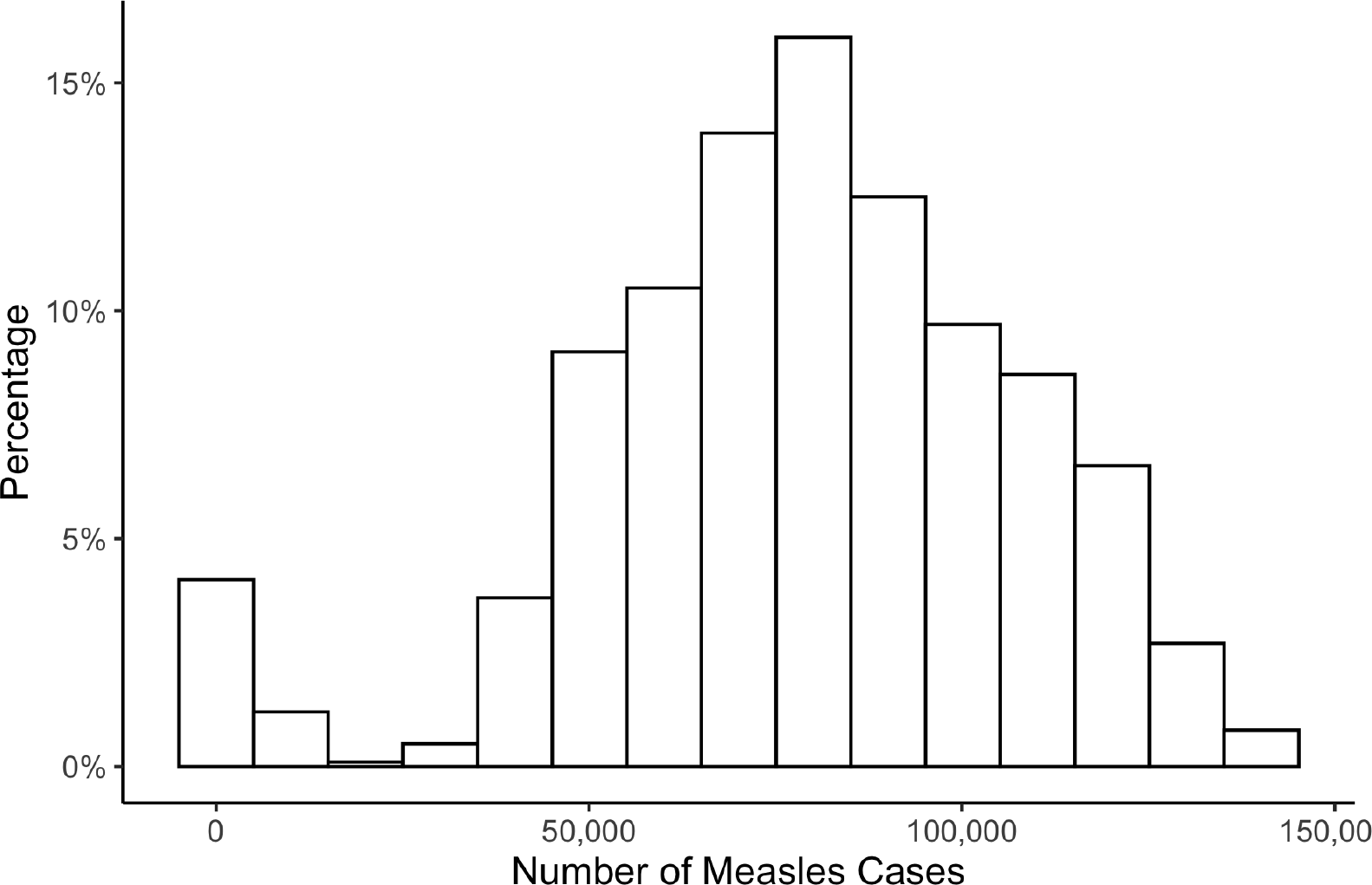
Histogram of estimated size of 2017 measles epidemic if vaccination had not occurred using LHS with 1,000 parameter combinations. Summary statistics: 78,151 (mean), 79,404 (median), 609 (2.5%), 127,147 (97.5%)

**Figure 3:**
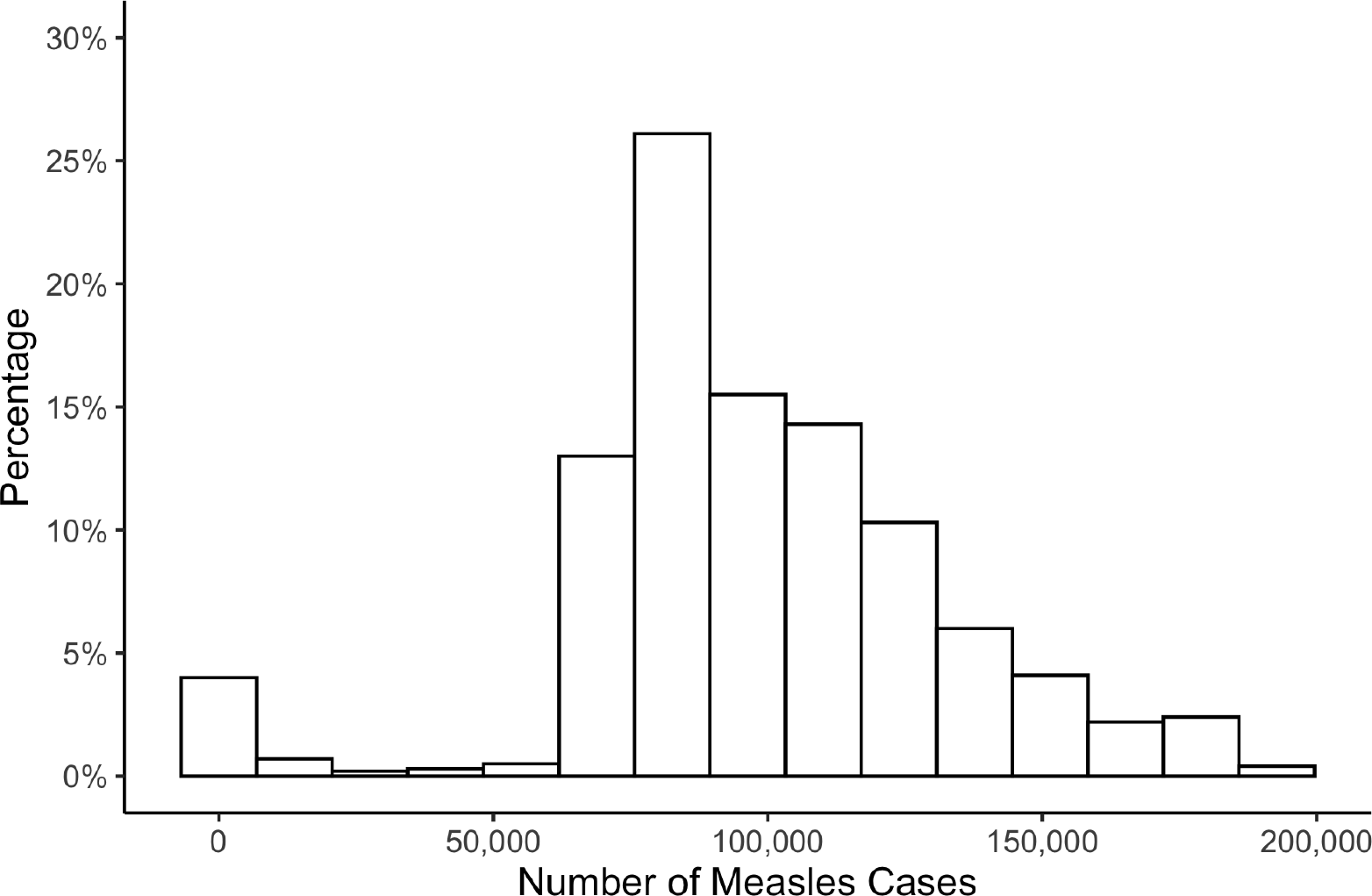
Histogram of estimated size of next measles epidemic if vaccination does not occur using LHS with 1,000 parameter combinations. Summary statistics: 97,637 (mean), 93,718 (median), 47 (2.5%), 173,397 (97.5%)

